# Characterization of External Defibrillator Output and Its Impact on Defibrillation Protection of Medical Equipment

**DOI:** 10.64898/2025.12.21.25342795

**Authors:** Dian Zhang, Jin Deng, Yong Yin

**Affiliations:** Shanghai Medical Device Services Certification & Testing, SGS-CSTC Standards Technical Services (Shanghai) Co., Ltd., Shanghai 201612, PR China

**Keywords:** Cardiac arrest, Resuscitation, Biphasic waveforms, Defibrillation voltage, Defibrillation energy

## Abstract

**BACKGROUND:** Defibrillators constitute a critical component of emergency care for ventricular fibrillation and pulseless ventricular tachycardia. Advances in defibrillation technology point to the potential need for reassessing current defibrillation protection standards and associated resuscitation guidelines.

**OBJECTIVE:** This study aimed to explore potential implications for future defibrillator requirements, defibrillation protection standards, and resuscitation guidelines based on the parameters of commercially available defibrillators.

**METHODS:** Defibrillator brands were identified from the registration records of the National Medical Products Administration, the U.S. Food and Drug Administration, and the European Database on Medical Devices. Key defibrillation parameters were collected from manufacturer-provided data.

**RESULTS:** All studied devices employ biphasic waveforms, with biphasic truncated exponential (BTE) being the most common, followed by rectilinear biphasic (RB), and a smaller number using pulsed biphasic (PB). Except for PB devices, maximum output voltages are below 3 kV. While energy required for effective defibrillation decreased, some manufacturers retain an upper limit of 360 J.

**CONCLUSION:** Incorporating biphasic waveforms into defibrillation protection tests may be appropriate. Adjustments to the requirements and guidelines of defibrillation voltage or energy could be considered, depending on market trends and future studies on efficacy.

## 1 Introduction

Electrical defibrillation is effective and essential in the emergency management of sudden cardiac arrest caused by malignant arrhythmias, such as ventricular fibrillation and pulseless ventricular tachycardia.^1,2^ A strong shock which induces sufficient myocardial depolarization can terminate these arrhythmias and restore sinus rhythm.^1^ The first direct current defibrillator was developed to deliver such shocks by Naum Gurvich in 1939.^3^ However, shocks delivered by external defibrillators may interfere with or damage other medical devices when used concurrently. This risk prompted the development of defibrillation-proof standards. In the last century, monophasic waveforms were widely employed, such as damped sine waves, among early defibrillators. Consequently, standards related to defibrillation protection were primarily established based on the performance specifications of monophasic devices. When the era arrived at the beginning of 21st century, biphasic defibrillators with lower defibrillation energy gradually replaced them.^5^ As a result, the corresponding resuscitation guidelines and standards may require revision. The aim of this study was to investigate the performance parameters of external defibrillators currently available on the market and to assess their potential impact on defibrillation protection.

## 2 Methods and Materials

Defibrillator brands were identified from valid registration records in the medical device databases of the National Medical Products Administration, the U.S. Food and Drug Administration, and the European Database on Medical Devices, using the keywords “defibrillator” and “defibrillation”. Accessories, such as single-use defibrillator pads, were excluded first. As this study focused on external defibrillation, implantable cardioverter defibrillators were subsequently excluded. There brands (SUPERSTAR MED, Sinopharm Group Co. Ltd, and Creative Medical) were also excluded due to limited public information. Therefore, the final list comprised 31 manufacturers (see Table 1). In this study, products were categorized into three groups based on the intended use, which are automated external defibrillators (AEDs), defibrillator monitors, and wearable cardioverter defibrillators (WCDs), respectively. Technical parameters, including operating mode, waveform, defibrillation energy, output voltage, output current, and patient impedance, were extracted from product manuals, brochures, official websites, registration databases, and third-party distributors, with data current as of August 25, 2025.

**Table 1.**
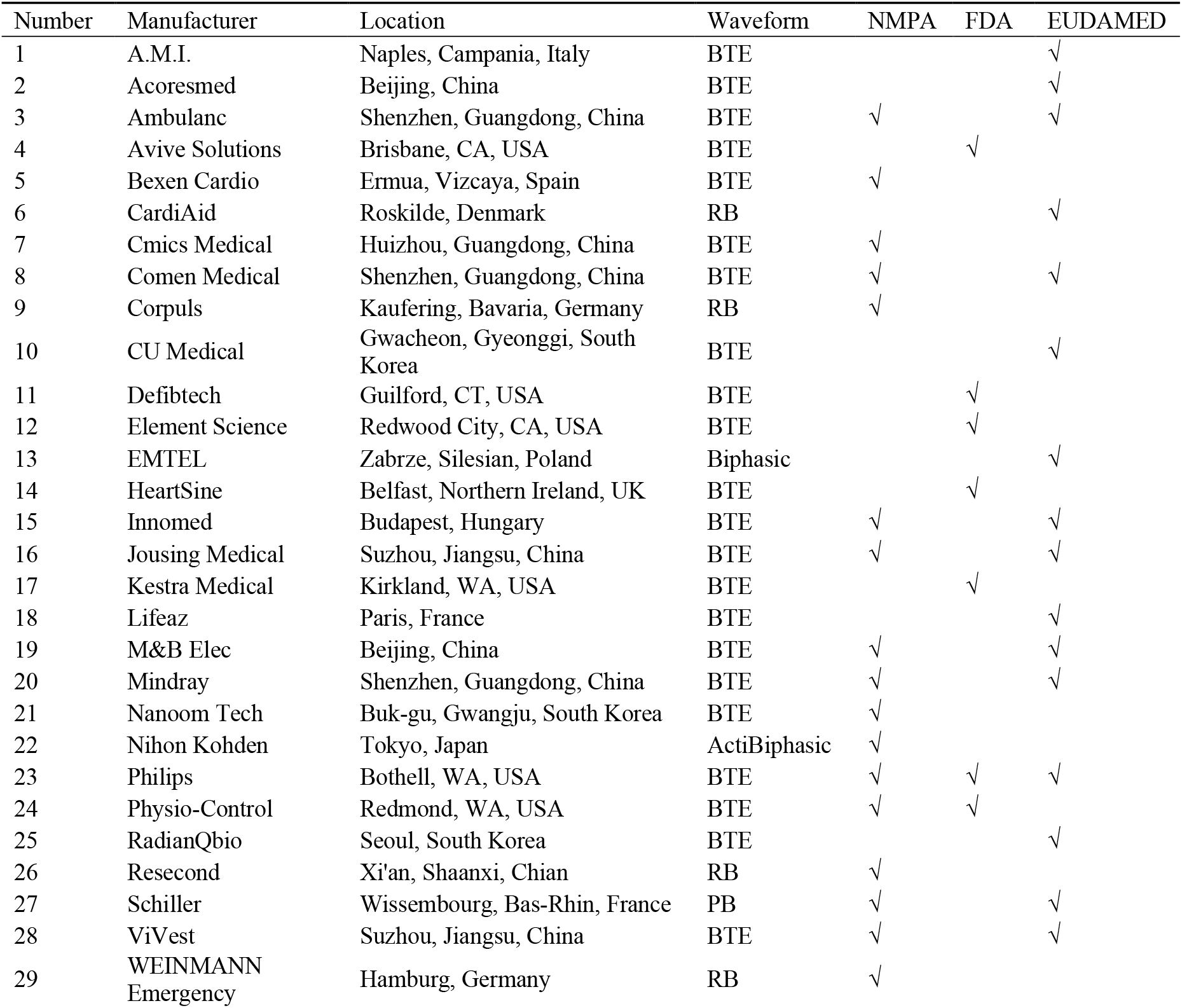
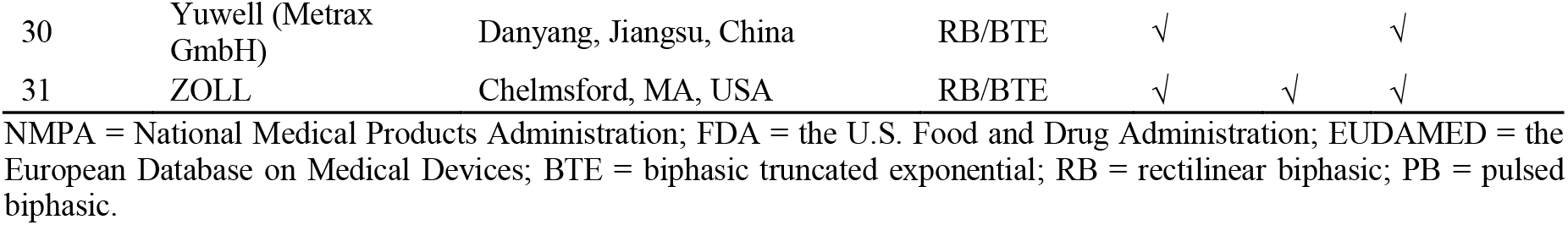
Defibrillator manufacturers and their products’ defibrillation waveforms.

| Number | Manufacturer | Location | Waveform | NMPA | FDA | EUDAMED |
| --- | --- | --- | --- | --- | --- | --- |
| 1 | A.M.I. | Naples, Campania, Italy | BTE |  |  | √ |
| 2 | Acoresmed | Beijing, China | BTE |  |  | √ |
| 3 | Ambulanc | Shenzhen, Guangdong, China | BTE | √ |  | √ |
| 4 | Avive Solutions | Brisbane, CA, USA | BTE |  | √ |  |
| 5 | Bexen Cardio | Ermua, Vizcaya, Spain | BTE | √ |  |  |
| 6 | CardiAid | Roskilde, Denmark | RB |  |  | √ |
| 7 | Cmics Medical | Huizhou, Guangdong, China | BTE | √ |  |  |
| 8 | Comen Medical | Shenzhen, Guangdong, China | BTE | √ |  | √ |
| 9 | Corpuls | Kaufering, Bavaria, Germany | RB | √ |  |  |
| 10 | CU Medical | Gwacheon, Gyeonggi, South Korea | BTE |  |  | √ |
| 11 | Defibtech | Guilford, CT, USA | BTE |  | √ |  |
| 12 | Element Science | Redwood City, CA, USA | BTE |  | √ |  |
| 13 | EMTEL | Zabrze, Silesian, Poland | Biphasic |  |  | √ |
| 14 | HeartSine | Belfast, Northern Ireland, UK | BTE |  | √ |  |
| 15 | Innomed | Budapest, Hungary | BTE | √ |  | √ |
| 16 | Jousing Medical | Suzhou, Jiangsu, China | BTE | √ |  | √ |
| 17 | Kestra Medical | Kirkland, WA, USA | BTE |  | √ |  |
| 18 | Lifeaz | Paris, France | BTE |  |  | √ |
| 19 | M&B Elec | Beijing, China | BTE | √ |  | √ |
| 20 | Mindray | Shenzhen, Guangdong, China | BTE | √ |  | √ |
| 21 | Nanoom Tech | Buk-gu, Gwangju, South Korea | BTE | √ |  |  |
| 22 | Nihon Kohden | Tokyo, Japan | ActiBiphasic | √ |  |  |
| 23 | Philips | Bothell, WA, USA | BTE | √ | √ | √ |
| 24 | Physio-Control | Redmond, WA, USA | BTE | √ | √ |  |
| 25 | RadianQbio | Seoul, South Korea | BTE |  |  | √ |
| 26 | Resecond | Xi'an, Shaanxi, Chian | RB | √ |  |  |
| 27 | Schiller | Wissembourg, Bas-Rhin, France | PB | √ |  | √ |
| 28 | ViVest | Suzhou, Jiangsu, China | BTE | √ |  | √ |
| 29 | WEINMANN<br>Emergency | Hamburg, Germany | RB | √ |  |  |
| 30 | Yuwell (Metrax GmbH) | Danyang, Jiangsu, China | RB/BTE | √ |  | √ |
| 31 | ZOLL | Chelmsford, MA, USA | RB/BTE | √ | √ | √ |

NMPA = National Medical Products Administration; FDA = the U.S. Food and Drug Administration; EUDAMED = the European Database on Medical Devices; BTE = biphasic truncated exponential; RB = rectilinear biphasic; PB = pulsed biphasic.

If the documentation specifies the peak or maximum voltage of a defibrillation pulse, it is taken as the maximum output voltage (V_max_). Otherwise, according to the International Electrotechnical Committee (IEC) standards 60601-2-4 (Clause 201.7.9.3.101), manufacturers must provide the time–voltage or time–current waveform^,4^ enabling the determination of the maximum voltage. When only the peak current is reported, the peak voltage is derived from Ohm’s law, *U* = *IR*, where *I* is the peak current and *R* is the patient impedance. For biphasic truncated exponential (BTE) waveforms, the maximum current occurs at the first-phase peak, as in most other biphasic waveforms. ZOLL data indicate that at low impedances, the second-phase current can exceed the first. Therefore, for rectilinear biphasic (RB) waveforms, the maximum initial currents of the first and second phases must be compared. In this study, V_max_ was consistently observed in the first phase of the defibrillation pulse. If waveform diagrams lacked numerical values, the peak current or voltage was estimated from the scales.

## 3 Results

### 3.1 Operating Mode

From the perspective of operating mechanism, AEDs can be classified as semi-automatic or fully automatic. Defibrillator monitors generally support manual defibrillation and AED modes. The three WCDs—ZOLL LifeVest, Kestra Medical ASSURE WCD System, and Element Science Jewel—use fully automatic defibrillation. To accommodate age diversity, most AEDs offer adult and pediatric modes, with some limited to adults.

### 3.2 Defibrillation Waveform

The results as investigated identified that all devices with publicly available information employ biphasic waveform defibrillation (see Table 1). Within all valid results, EMTEL only states that its DefiMax defibrillator monitors use a biphasic waveform, without detailing the waveshape. Among the remaining manufacturers, BTE waveforms are most common (see Figure 1(a)), appearing in products from 24 of 31 manufacturers, including notable brands such as Philips and Physio-Control. Nihon Kohden’s ActiBiphasic is an exception, featuring a truncated exponential waveform in the first phase and a power-controlled waveform in the second (see Figure 1(b)). As for ZOLL products, the LifeVest uses BTE waveforms, whereas AEDs (AED Plus and its fully automatic version, ZOLL AED 3, and AED Pro) and defibrillator monitors (X and R series) employ rectilinear biphasic (RB) waveforms (see Figure 1(c)). Resecond reports that the RB waveform in its AEDs (RCD-500, RCD-600, and RCD-600E) is equivalent to the ZOLL waveform. Additionally, the Corpuls3 series defibrillator monitors from Corpuls also employ RB waveforms. WEINMANN Emergency designed a sawtooth-shaped pulse, with similar waveforms in CardiAid devices, both treated as RB in this study. Metrax GmbH (part of the Chinese Yuwell Group since 2017) uses BTE in the HeartSave Y series and RB in both the HeartSave AED series (M250) and Xdxe defibrillator monitors. Except for ZOLL and Metrax GmbH, each manufacturer typically employs a single waveform type across its products. Pulsed biphasic (PB) waveforms are less common. Only Schiller’s FRED easy AED and DEFIGARD 5000 defibrillator monitors use a PB waveform, termed “Multipulse Biowave” (see Figure 1(d)).

**Figure 1.**
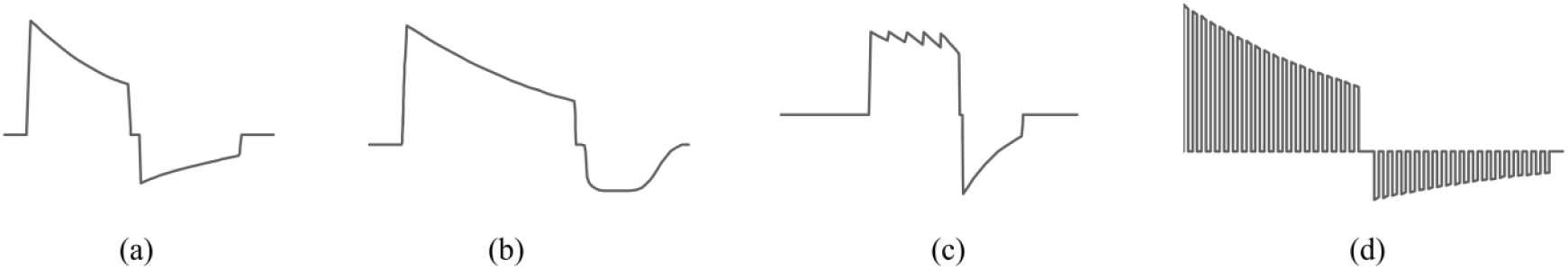
Biphasic defibrillation waveforms. (a) biphasic truncated exponential waveform (Philips); (b) ActiBiphasic waveform (Nihon Kohden); (c) rectilinear biphasic waveform (ZOLL); (d) pulsed biphasic waveform (Schiller).

### 3.3 Defibrillation Energy

In adult and pediatric modes, common defibrillation energies are 150 J and 50 J, respectively (at 50 Ω). Some AEDs or AED modes in defibrillator monitors use fixed energy outputs, such as Philips HeartStart FRx and FR3, and ViVest PowerBeat X and M series. Among WCDs, the ASSURE WCD System delivers a fixed 170 J. Most AEDs escalate energy with successive shocks. For example, Nihon Kohden AEDs deliver 150–200–200 J (corresponding to the first, second, and third shocks, with the second and third shocks both at 200 J) in adult mode and 50–70–70 J in pediatric mode (second-shock escalation), M&B Elec AED 7000 uses 150–150–200 J (third-shock escalation), and ZOLL AEDs deliver 120–150–200 J in adult mode and 50–70–85 J in pediatric mode (gradual increase).

According to IEC 60601-2-4 (Clause 201.12.4.1), selectable defibrillation energy shall not exceed 360 J.^4^ Historically, the limit was established based on earlier monophasic waveforms. As biphasic waveforms require less energy to achieve effective defibrillation^,6,7^ manufacturers have taken different approaches when setting the maximum energy of their devices. For instance, WEINMANN Emergency limits the output of defibrillator monitors to 200 J, ZOLL applied a uniform 200 J limit across both AEDs and monitors, and Nihon Kohden applies 270 J for monitors but restricts AEDs for 200 J. By contrast, brands such as Physio-Control, Innomed, EMTEL, Mindray, and Comen Medical continue to support the traditional 360 J maximum—a choice to accommodate extreme conditions like high-impedance patients.

### 3.4 Maximum Output Voltage (V_max_)

Maximum output voltage was determined by comparing peak voltages across different patient impedances. Table 2, Table 3, and Table 4 present the V_max_ for AEDs, defibrillator monitors, and WCDs, respectively. This section includes 16 brands and 33 devices: 18 AEDs, 13 defibrillator monitors, and two WCDs. For voltages calculated from current, the corresponding peak current, patient impedance, and energy (selected or delivered, depending on manufacturer notation) are indicated. All other values were extracted from original sources.

**Table 2.**
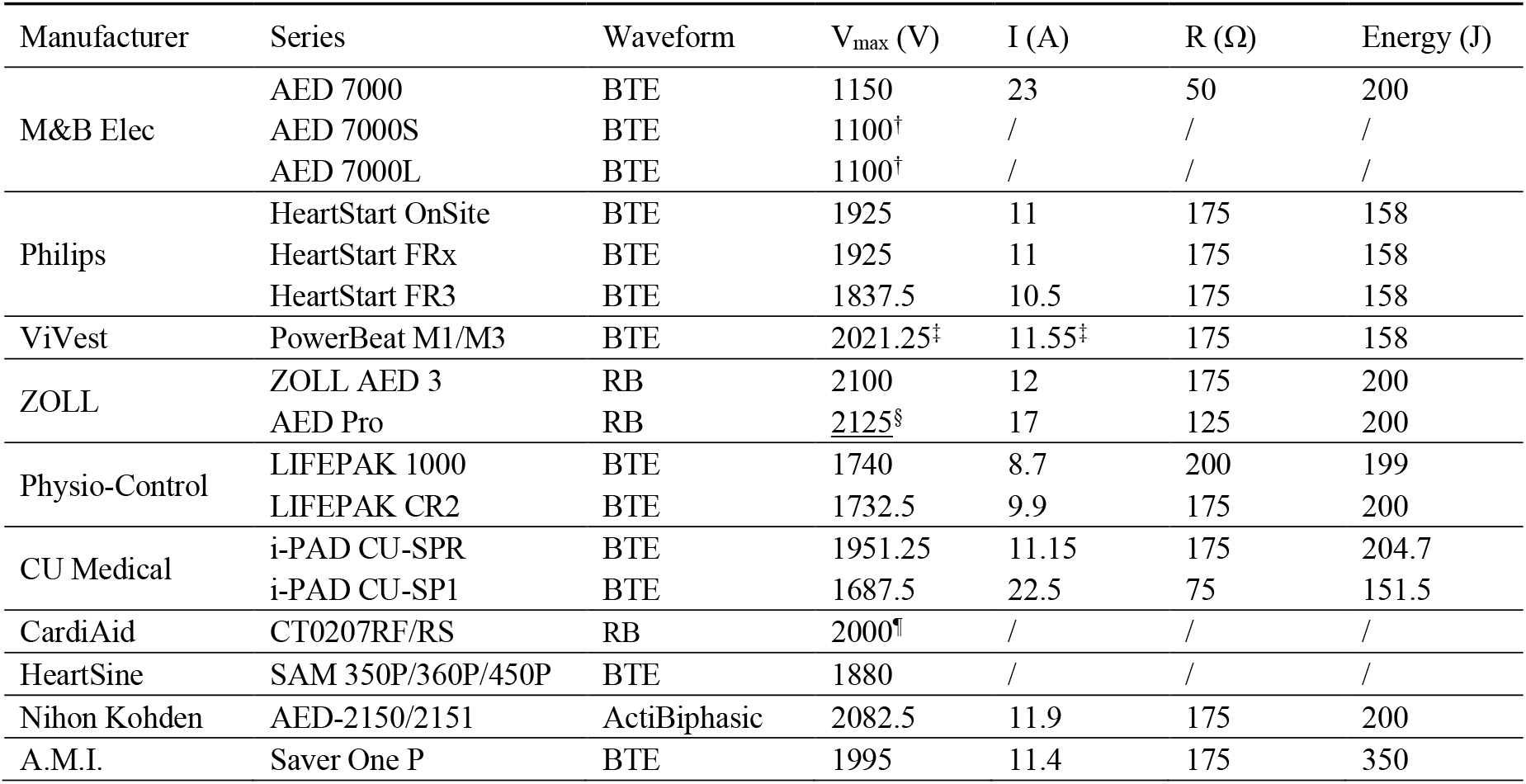
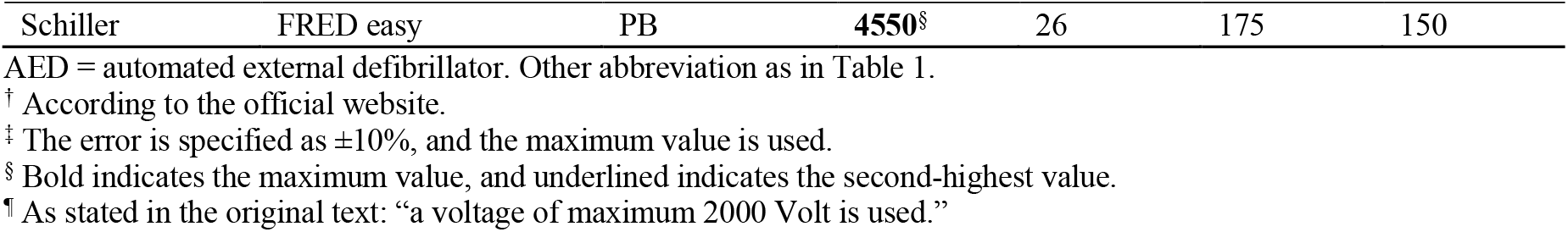
Maximum output voltage of AEDs.

**Table 3.**
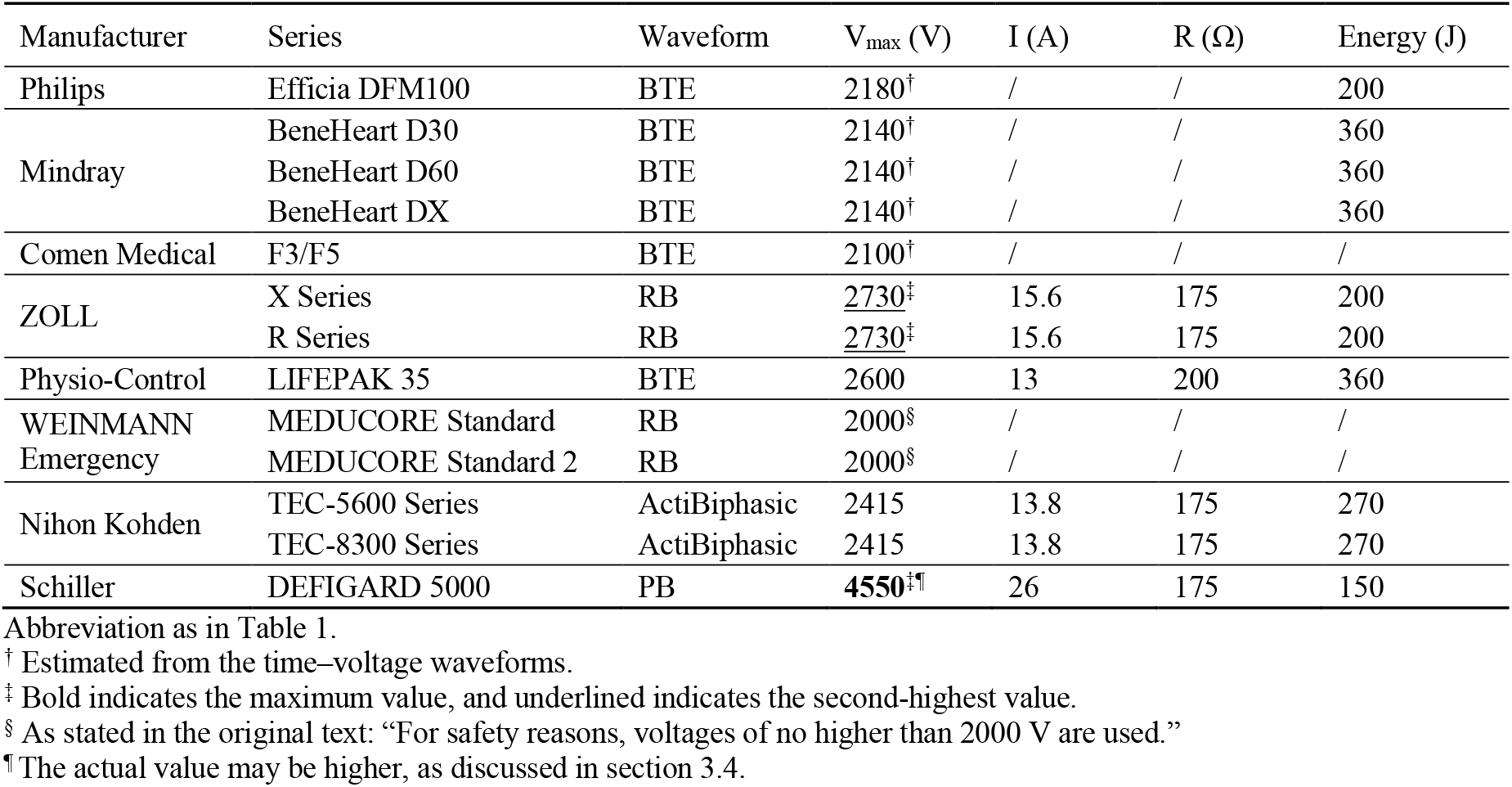
Maximum output voltage of defibrillator monitors.

**Table 4.** Maximum output voltage of WCDs.

| Manufacturer | Series | Waveform | $V_{\max}$ (V) | I (A) | R ($\Omega$) | Energy (J) |
| --- | --- | --- | --- | --- | --- | --- |
| Kestra Medical | ASSURE WCD System | BTE | 1650 | 11 | 150 | 126–148 |
| Element Science | Jewel | BTE | 1520 | 7.6 | 200 | 150/162 |

WCD = wearable cardioverter defibrillator. Other abbreviation as in Table 1.

As claimed above that Schiller products employ PB waveforms. Due to their waveform characteristics, V_max_ values are much higher than other defibrillators. Therefore, we temporarily exclude them and will discuss this phenomenon later. For the majority of products under survey, the V_max_ does not exceed 2 kV, or only slightly exceeds it. As for AEDs, 13 of 18 devices, including Philips HeartStart and Physio-Control LIFEPAK series, have a V_max_ below 2 kV. The ZOLL AED Pro exhibits the highest V_max_ in this category at 2125 V (125 Ω load). Defibrillator monitors generally exhibit higher V_max_ due to greater maximum delivered energy, with half number of products reporting V_max_ not exceeding 2.2 kV. Exceptions include ZOLL X and R series (2730 V), Physio-Control LIFEPAK 35 (2600 V at 200 Ω), and Nihon Kohden TEC-5600 and TEC-8300 series (2415 V). Furthermore, compared to others, the ASSURE WCD System and Jewel reach V_max_ of 1650 V (150 Ω) and 1520 V (200 Ω), respectively. Some products claim compatibility with patient impedances above 200 Ω. However, defibrillation waveform parameters reported by manufacturers only cover 25 to 175 Ω in 25-Ω intervals, consistent with IEC 60601-2-4 (Clause 201.7.9.3.101) requirement,^4^ with few extending to 200 Ω. Considering V_max_ is generally achieved at high-impedance loads, and some technical documentation reports theoretical values without measurement error, the actual V_max_ may be slightly higher, but remains well below the 5 kV test voltage for defibrillation protection.

Although Schiller’s Multipulse Biowave is biphasic, its V_max_ greatly exceeds comparable products. The FRED easy AED and DEFIGARD 5000 defibrillator monitors reach 4550 V at 175 Ω, corresponding to 150 J. Furthermore, the DEFIGARD 5000’s maximum energy is 180 J. As Zelinka et al. reported, the Schiller FRED achieves a peak current of 37 A at 180 J and 130 Ω,^8^ corresponding to 4810 V. Thus, the DEFIGARD 5000’s V_max_ at 175 Ω may approach 5 kV. In summary, excluding Schiller products, all commercially available defibrillators surveyed exhibit V_max_ below 3 kV.

## 4 Discussion

### 4.1 Waveform of defibrillation testing

Biphasic waveforms have been reported to exhibit comparable or superior efficacy compared with monophasic waveforms ^6,7,9,10^ while requiring lower energy, which facilitates the production of smaller devices. Some manufacturers have further reduced device size through innovative circuit designs. For instance, while phase inversion is typically achieved using an H-bridge circuit,^11^ Nihon Kohden has developed a design in which the inductor and capacitor discharge sequentially during the negative phase, generating a power-controlled waveform and eliminating the need for conventional H-bridge circuitry. In addition to BTE, RB, and PB waveforms, other waveforms, such as ascending-ramp biphasic waveforms^12^ and nanosecond electric shocks^,13^ have been proposed, although none have been applied in practice. The 2010 European Resuscitation Council (ERC) guidelines note that monophasic defibrillators are no longer manufactured^,5^ and this edition represents the last ERC guideline to mention monophasic waveforms. Notably, Physio-Control’s LIFEPAK 12 was available in both monophasic and biphasic versions, with the monophasic version discontinued in 2013. Based on our results, the most widely used biphasic waveforms are BTE and RB. Their relative efficacy remains debated, with some studies suggesting comparable performance.^14^ RB waveforms maintain high current even at high impedance, ensuring effective defibrillation. As a result, their maximum output voltage is relatively higher, which will be further discussed in section 4.3.

Despite years of advancement in biphasic waveforms, current standards continue to use monophasic waveforms for defibrillation protection tests, reflecting a conservative worst-case approach. However, an animal study in dogs found that when the second phase of a biphasic waveform exceeded the first, the defibrillation threshold was higher than that of a monophasic waveform of equivalent duration.^15^ From this case, we concluded that using test circuits producing a monophasic waveform cannot fully represent the worst-case. Therefore, adopting a strategy similar to the standard designed for active implantable medical devices (ISO 14708-1 Clause 20.2), which tests monophasic and biphasic waveforms separately,^16^ may be a reasonable approach during the transitional period when the residual use of monophasic waveforms is uncertain. Once sufficient evidence supports the complete phase-out of monophasic waveforms, tests can then evolve to include a representative set of biphasic waveforms.

### 4.2 Defibrillation energy limit

The primary advantage of biphasic over monophasic defibrillators is the reduced energy required for defibrillation, but as mentioned in section 3.3, some manufacturers maintain an upper energy limit of 360 J. At present, the guidance on biphasic defibrillation energy is fairly broad and is largely determined by manufacturer-specified doses. The ERC guidelines 2021^17^ recommend 150–360 J for the second and subsequent shocks, with an upper limit of 360 J, whereas the 2020 American Heart Association guidelines^2^ do not specify a maximum energy limit for biphasic defibrillation. In the previous studies, higher biphasic energy levels (360 J) are not more harmful than lower ones (150 J) in the swine model,^18^ and patients in ventricular fibrillation benefit from higher energy biphasic defibrillation in multiple shocks.^19^ Therefore, 360 J biphasic defibrillation is acceptable for non-first shock.

However, the efficacy of higher energy biphasic defibrillation in humans is debated. While an escalating-energy strategy (200–300–360 J) performs better than a fixed low energy defibrillation in both animal models (150 J) and humans (200 J)^,20,21^ there is no better survival rate at 30 days in humans when comparing the escalating-energy and fixed high energy (360 J) defibrillation.^22^ Double sequential defibrillation is a resuscitation strategy in which two defibrillators with different pad placement (anterior-laterally and anterior-posterior) discharge almost simultaneously. It was identified effective in some case reports,^23,24,25^ although there are not confirmed association between double sequential defibrillation and enhanced outcomes.^26,27,28^ In this case, the delivered energy (more than 400 J) is twice that of the standard defibrillation strategy, which potentially proved the efficacy of biphasic defibrillation with higher energy. As the relationship between energy levels and the efficacy and safety of biphasic defibrillation is still inconclusive, further study using different biphasic waveforms is needed.

### 4.3 Lower defibrillation voltage

According to IEC 60601-2-4 (Clause 201.12.4.101), the output voltage of a defibrillator must not exceed 5 kV when measured at a 175 Ω load.^4^ This limit is consistent with the 5 kV direct current voltage specified in the defibrillation protection test circuit of IEC 60601-1 (Clause 8.5.5.1 Figure 9 and Figure 10),^29^ as well as the term “Full no-load defibrillator voltage” mentioned in IEC 60601-2-25 (Subclause 201.8.5.5.1 Table AA.1).^30^ IEC 60601-2-25 (Subclause 201.8.5.5.1) further specifies that, when a medical device is positioned near the thorax or at a point electrically between the defibrillator paddles, the voltage should be slightly more than half of the no-load defibrillator voltage.^30^ In particular requirements, only IEC 60601-2-2 (Clause 201.8.5.5) lowers the voltage requirement to 2 kV for high-frequency surgical equipment and accessories.^31^ Based on the findings of this study, the maximum output voltage of most defibrillators is approximately 2 kV. Except for Schiller products, the highest value recorded is 2730 V for the ZOLL X and R series, which remains below 3 kV even with a ±5% margin of error. Therefore, it may be considered to lower the defibrillation protection test voltage accordingly. Reducing the test voltage could shorten the required insulation distance. For example, if the defibrillation voltage is reduced from 5 kV to 3 kV, according to Table F.2 of IEC 60664-1, the required clearance distance in air up to 2 000 m above sea level can be adjusted to 2.0 mm (originally 4.0 mm) in the case of inhomogeneous field.^32^ This could make the design of medical devices more compact, which is particularly important for devices that need to operate in limited spaces, such as increasingly adopted WCDs, portable monitors or emergency equipment.

The PB waveform reduces the energy required for defibrillation through pulsed delivery, while maintaining a relatively high output voltage. It represents another direction in the development of biphasic waveform technology, showing greater advantages in reducing defibrillation energy compared with BTE and RB waveforms. Due to the diversity in the development of biphasic waveform technologies, the formulation of standards and resuscitation guidelines are influenced by the market. In the 2021 ERC guidelines, recommendations for defibrillation energy included the PB waveform for the first time,^17^ demonstrating its growing impact. This waveform does not offer an advantage in output voltage. However, if it demonstrates superior efficacy and safety compared with other biphasic waveforms in the future, updates to standards may prioritize lower recommended defibrillation energies. As suggested in section 4.1, multiple biphasic waveforms may be employed in future defibrillation testing. In line with this section, distinct voltage limits can further be specified for each waveform.

### 4.4 Limitation and future work

Due to resource limitations, this study relied on manufactured-specified parameters rather than measured ones. Future work may validate the findings across representative brands and models. A further limitation of this study is its focus on currently available products, excluding defibrillators that have been discontinued but may still be in use. Monophasic waveforms have been discontinued for several years, and their current usage is presumed to be limited. For instance, Physio-Control LIFEPAK 10 had its most recent recall notice in 2008, while LIFEPAK 12 monophasic was officially discontinued in 2013. Another concern is that some older biphasic models also have relatively high maximum output voltages. For example, the LIFEPAK 12 biphasic, with a V_max_ exceeding 3250 V,^8^ was discontinued in 2016, and maintenance support ceased in 2020. There may be other products with similar circumstances, though verifying them is challenging. However, with the continuous evolution of defibrillators, reducing the defibrillation protection voltage is promising and reasonable.

## 5 Conclusion

Through an investigation of the defibrillation parameters of commercially available devices, several trends in waveform usage and energy settings have been identified. All studied devices employ biphasic waveforms, with BTE being the most prevalent, followed by RB, and a smaller number using PB. Representative companies for each waveform type include Philips and Physio-Control for BTE, ZOLL for RB, and Schiller for PB. Except for PB devices, the maximum output voltages of defibrillators do not exceed 3 kV. Upper defibrillation energy is partially reduced in some devices, whereas others retain a maximum of 360 J. These characteristics influence future requirements for cardiac defibrillators, which in turn indirectly affect defibrillation protection standards. During the transitional period from monophasic to biphasic waveforms, test circuits may incorporate biphasic waveforms. Once monophasic waveforms are confirmed to be obsolete, they can be removed from the test protocols. The future direction of defibrillation protection of medical equipment is likely to be influenced by the market share of different manufacturers. For BTE or RB devices, standards could lower output voltage requirements, thereby relaxing defibrillation protection voltages. For PB devices, standards may reduce the recommended defibrillation energy. Looking further ahead, future biphasic waveform technologies are expected to provide high defibrillation efficacy while maintaining both lower voltage and lower energy, which has the potential to improve survival rates in sudden cardiac arrest.

## Data Availability

All data produced in the present study are available upon reasonable request to the authors

## Abbreviations

Abbreviation Definition

AEDs: Automated External Defibrillators
BTE: Biphasic Truncated Exponential
ERC: European Resuscitation Council
IEC: International Electrotechnical Committee
PB: Pulsed Biphasic
RB: Rectilinear Biphasic
Vmax: Maximum Output Voltage
WCDs: Wearable Cardioverter Defibrillators

